# Diurnal Heart Rate Range and Heart Rate Excursions: Novel Characterizations of Wearable Heart Rate Variability With an Application to Aging

**DOI:** 10.64898/2026.09.29.26364190

**Authors:** Samuel D. Fansler, Owen Yoo, Shuiqing Han, Luigi Ferrucci, Eleanor M. Simonsick, Jennifer A. Schrack, Irina Gaynanova, Vadim Zipunnikov

## Abstract

**Background:** Wearable heart rate (HR) monitors collect minute-level HR continuously across days, yet these data are commonly reduced to scalars such as resting HR (RHR) and heart rate reserve (HRR), defined as the difference between maximum HR and RHR. This discards two dimensions of within-day HR dynamics: how HR range varies across the day and the magnitude and duration of individual HR rises and declines. We introduce diurnal heart rate range (dHRR), a time-of-day-specific measure of HR range across repeated days, and heart rate excursions (HREs), individual HR rises and declines characterized by amplitude and duration.

**Methods:** We analyzed minute-level HR from 750 participants in the Baltimore Longitudinal Study of Aging (mean [SD] age, 66 [12] years). We estimated the 5th, 50th, and 95th HR percentiles at each minute of the 24-hour cycle across repeated days, defining diurnal HR (dHR) as the median curve and dHRR as the 95th minus 5th percentile curve. A moving-average algorithm segmented HR into HREs. Function-on-scalar regression modeled dHR and dHRR by age, sex,and BMI across the day; local polynomial regression examined age and sex differences in HRR and HREs.

**Results:** Conventional HRR decreased with age, indicating a smaller overall observed HR range. Age-related differences in dHR varied substantially across the day: between ages 40 and 80, fitted dHR was approximately 2 bpm lower at 3:00 AM but 8 bpm lower at 6:00 PM. dHRR showed that this contraction was not uniform across the day: the evening peak near 6:00 PM at younger ages was largely absent by age 70, while the morning peak shifted from approximately 9:00 AM toward noon. At the event level, HREs showed two parallel age-related changes: between ages 40 and 80, mean excursion amplitude decreased from approximately 40 to 32 bpm, while mean excursion duration increased from approximately 90 to 104 minutes.

**Conclusions:** Aging was characterized by a smaller global HR range, compression and reorganization of HR range across the day, and smaller, longer-lasting HREs. dHRR and HREs provide complementary views of the diurnal organization and event-level dynamics of wearable HR and are implemented in the open-source *ihr* R package.

## 1. Introduction

Wearable heart rate (HR) sensors have become increasingly common in everyday use and large-scale research studies, providing continuous streams of HR data under free-living conditions over days and weeks. Large initiatives such as the All of Us Research Program^1^ and the Adolescent Brain Cognitive Development Study^2^ have demonstrated the feasibility of collecting minute-level HR data from thousands of participants. These data provide an opportunity to characterize HR variability during daily life at a level of detail that is not available from conventional clinical measurements.

Despite this richness, wearable HR data are commonly reduced to a small number of scalar summaries, including resting HR (RHR), maximum HR, and heart rate reserve (HRR).^3^ These measures characterize overall HR level and global range but discard two fundamental dimensions of within-day HR dynamics, illustrated across the three panels of Figure 1. Panel A shows the conventional landmark representation, in which HRR summarizes the global range between RHR and maximum HR. Panel B extends this representation across the 24-hour day through diurnal heart rate range (dHRR), which describes how the repeatedly observed HR range expands and contracts with clock time. Panel C focuses on an individual heart rate excursion (HRE), a rise and subsequent decline in HR that can be characterized by its amplitude and duration.

**Figure 1.**
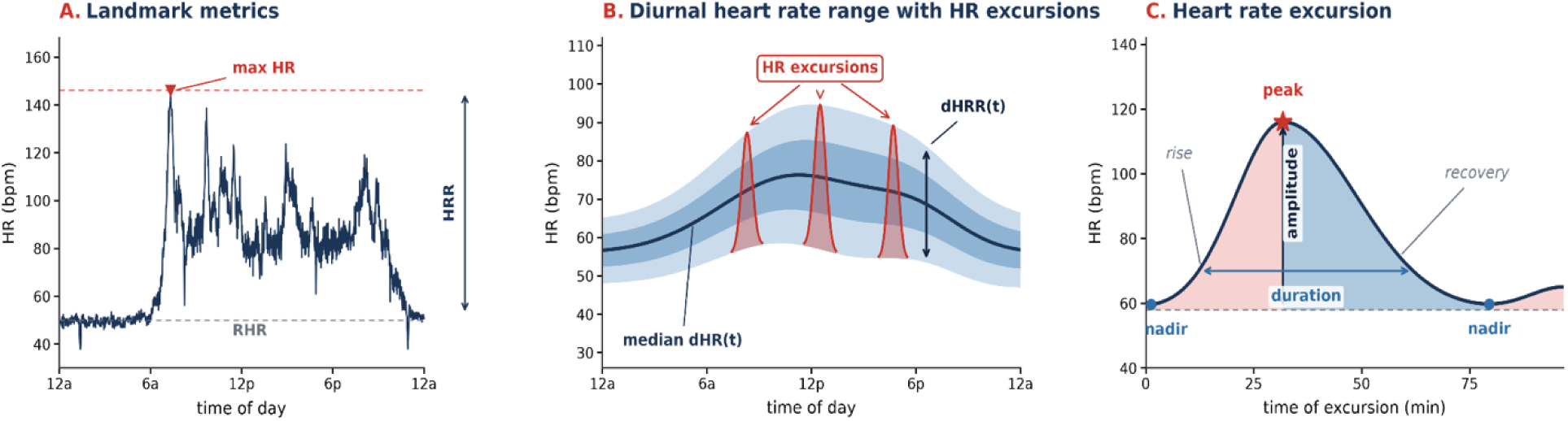
Conceptual overview of the three complimentary temporal representations derived from the same minute-level wearable HR signal. A: landmark metrics (RHR, maximum HR, and HRR) reduce the recording to global scalar values. B: the diurnal representation summarizes the median HR curve (dHR) and the time-of-day-specific 5th-to-95th percentile range (dHRR) across repeated days. Example individual HR excursions are illustrated. C: heart rate excursions (HREs) characterize individual localized rises and declines by amplitude and duration.

Figure 2 illustrates this loss of information using approximately one week of data from a participant in the Baltimore Longitudinal Study of Aging (BLSA).^4^ The minute-level trajectories contain thousands of observations, whereas conventional RHR and HRR compress the recording to a small number of landmarks. In contrast, the diurnal representation summarizes the distribution of HR repeatedly observed at each clock time and therefore shows when the observed HR range expands and contracts throughout the day.

**Figure 2.**
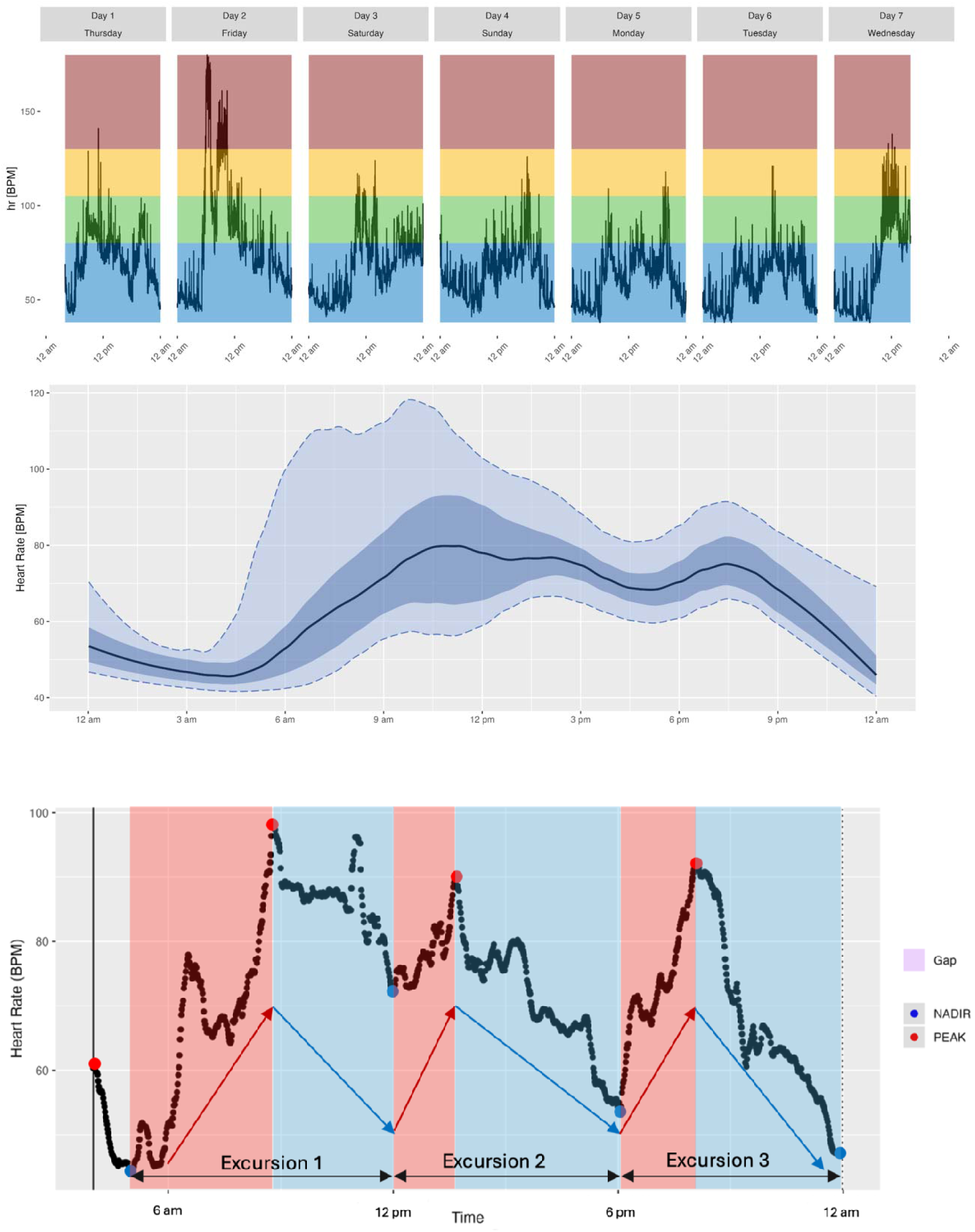
A HR data from seven days from a participant from the Baltimore Longitudinal Study of Aging. **Top panel:** raw minute-level HR from approximately seven days of device wear. Colors correspond to participant-level physical activity thresholds. **Middle panel:** estimated diurnal quantile curves. The black line represents median dHR, the darker band represents the diurnal interquartile range, and the lighter band represents the 5th-to-95th percentile typical daily HR envelope. **Bottom panel:** Three heart rate excursions: red regions correspond to rising HR portions ending in detected peaks; blue regions correspond to subsequent descending portions ending in nadirs.

We therefore propose dHRR and HREs as two complementary approaches to retaining information that conventional scalar summaries discard. dHRR captures how the observed HR range varies across the day, whereas HREs capture individual HR rises and declines and allow their amplitude and duration to be quantified.

A similar distinction between diurnal distributions and individual events has been useful in continuous glucose monitoring (CGM). The Ambulatory Glucose Profile, for example, summarizes multiple days of glucose readings using median and distributional curves across the 24-hour day,^5^ while measures of glycemic excursions characterize individual changes in glucose. Together with metrics such as time in, above, and below range, these complementary measures have been incorporated into international consensus recommendations and are widely used in clinical research and diabetes management.^6–10^ The CGM experience demonstrates how standardized representations of continuous sensor data can make information at different temporal scales interpretable and usable across research, clinical trials, and disease management. Similar principles can be applied to wearable HR, while recognizing that HR and glucose have different physiology and that HR-specific measures require independent evaluation.

Motivated by these considerations, this study makes three primary contributions. First, we introduce dHRR and HREs as complementary approaches to characterizing wearable HR variability, capturing the diurnal organization of HR range and the characteristics of individual HR changes, respectively. Second, we apply these approaches to BLSA data to examine how aging is associated with global HR range, its organization across the 24-hour day, and the amplitude and duration of individual HR excursions. Third, we integrate conventional landmark measures, diurnal distributions, and event-based measures into an Ambulatory Heart Rate Profile (AHP), with an open-source workflow implemented in the *ihr* R package.

## 2. New Heart Rate Measures

Wearable minute-level HR data can be represented at multiple temporal resolutions, with each scale addressing a different question.

### 2.1 Standard HR Landmark Metrics

In the conventional analysis framework, minute-level data are coarsened into scalar summaries using HR landmarks. Examples include RHR, which describes HR during a specified low-activity or resting period, and HRR, which describes the difference between a lower HR level and an observed maximum. Following Speed et al.,^11^ we define RHR as the average HR between 3:00 and 7:00 AM and conventional HRR as the difference between maximum observed HR and RHR. This measure is closely related conceptually to the long history of using the difference between resting and maximal HR to individualize exercise intensity.^12^ Within the physical activity literature, HRR is frequently used to construct individualized thresholds for physical activity intensities such as sedentary behavior, light activity, and moderate-to-vigorous activity.^3,13^ Each minute of HR can then be classified relative to the individual’s HRR-derived thresholds. These standard summaries are useful but discard temporal organization.

### 2.2 Diurnal Variability

The 24-hour circadian system regulates sleep, activity, energy, and numerous physiological processes. Disruptions in daily rhythmicity have been associated with adverse cardiovascular and other health outcomes.^14,15^ HR also follows a strong 24-hour pattern, with lower values during sleep and generally higher levels during waking activity.^16^ A single HRR computed over an entire monitoring interval cannot represent these pronounced time-of-day differences. We therefore derive subject-specific HR distributions as functions of clock time.

#### 2.2.1 Diurnal Heart Rate

For participant *i*, let *Q_i,p_(t)* denote the *p*th quantile of HR across repeated valid days at clock minute *t*. We define the participant’s diurnal HR curve as *dHR_i_(t)* = *Q_i,0.50_(t)*.

Thus, *dHR_i_(t)* is the median HR repeatedly observed from participant *i* at a particular clock time *t*. Unlike a single average HR, the dHR curve retains the overall shape of the daily HR, including the overnight nadir, morning increase, daytime plateau, and evening and nighttime declines.

#### 2.2.2 Diurnal Heart Rate Range

Around the median dHR curve, we estimate diurnal quantiles across repeated days. We define dHRR for participant *i* at time *t* as *dHRR_i_(t)* = *Q_i,0.95_(t)* – *Q_i,0.05_(t)*, where *Q_i,0.95_(t)* and *Q_i,0.05_(t)* are the 95th and 5th percentile HR values at clock time *t*. We refer to the region between these curves as the typical daily HR range. This definition differs from conventional HRR in several important ways.

First, dHRR is specific to the time-of-day. Conventional HRR returns one number, whereas dHRR is a function over 24 hours. It can therefore show that a person has a broad observed HR range in the morning but a narrow range overnight, or that range compression occurs preferentially during a particular part of the day. Second, dHRR is distribution-based and uses the repeated-day structure of wearable data. Rather than depending on a single observed maximum and one estimate of resting HR, it summarizes repeated observations using percentile boundaries at every clock time, transforming multiple days of minute-level measurements into a participant-specific function. This reduces dependence on isolated extreme observations while retaining a broad portion of the observed distribution. Third, dHRR preserves shape. The timing of morning expansion, presence or absence of an evening peak, nighttime compression, peak delay, and other aspects of the geometry of the daily HR envelope can themselves become outcomes of interest. Finally, because dHRR is a function, it can be analyzed using novel functional data methods that allow associations with age or other covariates to vary over the day instead of being represented by a single scalar coefficient.^17^

dHRR should not, however, be interpreted as a direct measure of maximal physiological or exercise-derived HR reserve. It is an observed free-living range and therefore reflects both cardiovascular responsiveness and the demands, behaviors, medications, environmental conditions, and other exposures present during the monitoring period. Its advantage is precisely that it quantifies this real-world range while retaining when that range occurs.

### 2.3 Heart Rate Excursions

Next, we define heart rate excursions (HREs) as localized rises and subsequent declines in HR occurring over minutes to hours (shown in the bottom panel of Figure 2). The detection strategy is adapted from excursion-detection approaches originally developed for continuous glucose monitoring (CGM) data.^18,19^ Short and long moving averages are used to smooth local noise and identify intervals likely to contain meaningful peaks and nadirs. Because this strategy was developed for CGM rather than HR, the smoothing windows and excursion thresholds should be viewed as tunable HR-analysis parameters rather than established physiological cut points.

For a qualifying excursion, we focus on two features: its amplitude, which reflects the magnitude of the HR response, and its duration, which reflects how long the elevation persists.

A central subject-level summary of amplitude used in this paper is the Mean Amplitude of Heart Rate Excursions (MAHE). MAHE is defined as the average amplitude of qualifying excursions exceeding the standard-deviation threshold used by the algorithm.

HRE amplitude can initially be interpreted as the magnitude of an observed free-living HR response, while HRE duration characterizes the persistence of the observed HR elevation or change. These constructs are related in motivation to laboratory measures of chronotropic response and post-exercise HR recovery, both of which have demonstrated prognostic relevance under standardized exercise conditions.^20–22^ However, free-living HREs are not equivalent to controlled exercise tests because the underlying stimulus, workload, posture, stress exposure, medication state, and environmental conditions are generally unknown.

Beyond amplitude and duration, each detected excursion also retains additional excursion-level features, including time to peak, recovery time, time of occurrence, and, across events, their frequency and the distributions of these quantities. Because the *ihr* package preserves the full event segmentation, these features are available as extensions of the framework (Supplementary Materials); in the present study, we focus on amplitude and duration.

Note that dHRR and HREs are complementary rather than interchangeable, and neither determines the other. Landmark measures such as RHR and maximum HR describe the global HR range; dHRR describes when the repeatedly observed range is broad or compressed; and HREs describe the individual trajectories that occur within that range. Together they separate four dimensions that conventional summaries can conflate: global range, diurnal organization, response magnitude, and persistence.

### 2.4 The *ihr* Package and the Ambulatory Heart Rate Profile

We provide the open-source R package Interpreting Heart Rate (*ihr*) to implement the proposed analytical framework directly from minute-level wearable HR data. The package computes conventional landmark summaries, diurnal quantile curves and dHRR, and HRE characteristics including MAHE. Additional functionality includes threshold-based HR episodes, rate-of-change measures, and other summaries described in the Supplementary Materials. The package is intended to make the proposed framework fully reproducible.

The Ambulatory Heart Rate Profile (AHP) provides a standardized participant-level summary of wearable HR data that integrates three complementary representations:

- **Minute-level HR:** HR observations across wear days.
- **Diurnal HR:** the median dHR curve, interquartile range, and 5th-to-95th percentile envelope.
- **Landmark summaries:** RHR, conventional HRR, maximum HR, and time spent in participant-specific HRR ranges.

## 3. Application to the Baltimore Longitudinal Study of Aging

### Data

The Baltimore Longitudinal Study of Aging, the longest-running longitudinal study of human aging, began in 1958.^4^ Minute-level HR monitoring using the Actiheart chest-worn monitor was introduced in 2007.^23^ For the present demonstration, we use BLSA data to illustrate the AHP, examine HR excursions for an individual participant, and evaluate cross-sectional associations between age and conventional, diurnal, and excursion-based HR measures. The analysis is intended as a proof-of-concept demonstration of the framework rather than an exhaustive evaluation or clinical validation of every proposed metric. For this analysis, we began with 890 participants aged 26 to 97 years who visited BLSA between 2007 and 2015. Because some participants had multiple visits, only the first qualifying visit was included. Participants older than age 85 were excluded because relatively few observations were available at these ages. A valid wear day was defined as a 24-hour period containing at least 90% wear time, corresponding to at least 1,296 of 1,440 possible minutes. Participants with fewer than three valid days were excluded. The final analytic sample included 750 individuals with mean age 66 years (SD = 12). Mean BMI was 27 kg/m² (SD = 5), and 51% of participants were female.

### Illustrative Example: Diurnal Profile and Excursions

Figure 2 shows the AHP for an example BLSA participant aged 66-70. The upper panel displays raw minute-level HR recorded across approximately seven days of continuous Actiheart wear. HR is visibly lower overnight and higher during waking periods, but considerable variation also occurs between days. The middle panel converts the repeated daily observations into clock-time-specific distributions. Median dHR increases in the morning, remains elevated through much of the daytime, shows a smaller later-day increase, and then declines during the night. The interquartile and 5th-to-95th percentile bands reveal that the amount of observed HR variation is itself strongly time dependent. The participant’s RHR is 55 bpm and maximum HR is 180 bpm, corresponding to a conventional HRR of 125 bpm. These scalar values describe the participant’s global low-to-high span, but they do not reveal the timing and shape of the repeated daily HR envelope shown in the middle panel. The bottom panel of Figure 2 illustrates three HREs during a 24-hour period for the same participant. Each detected excursion consists of an HR rise toward a peak followed by a subsequent decline toward a nadir. The events illustrate why retaining the excursion itself is useful.

## 4. Results

### Landmark-Based Metrics and Heart Rate Excursions

Figure 3 summarizes age associations for conventional landmark metrics and excursion-based measures.

**Figure 3.**
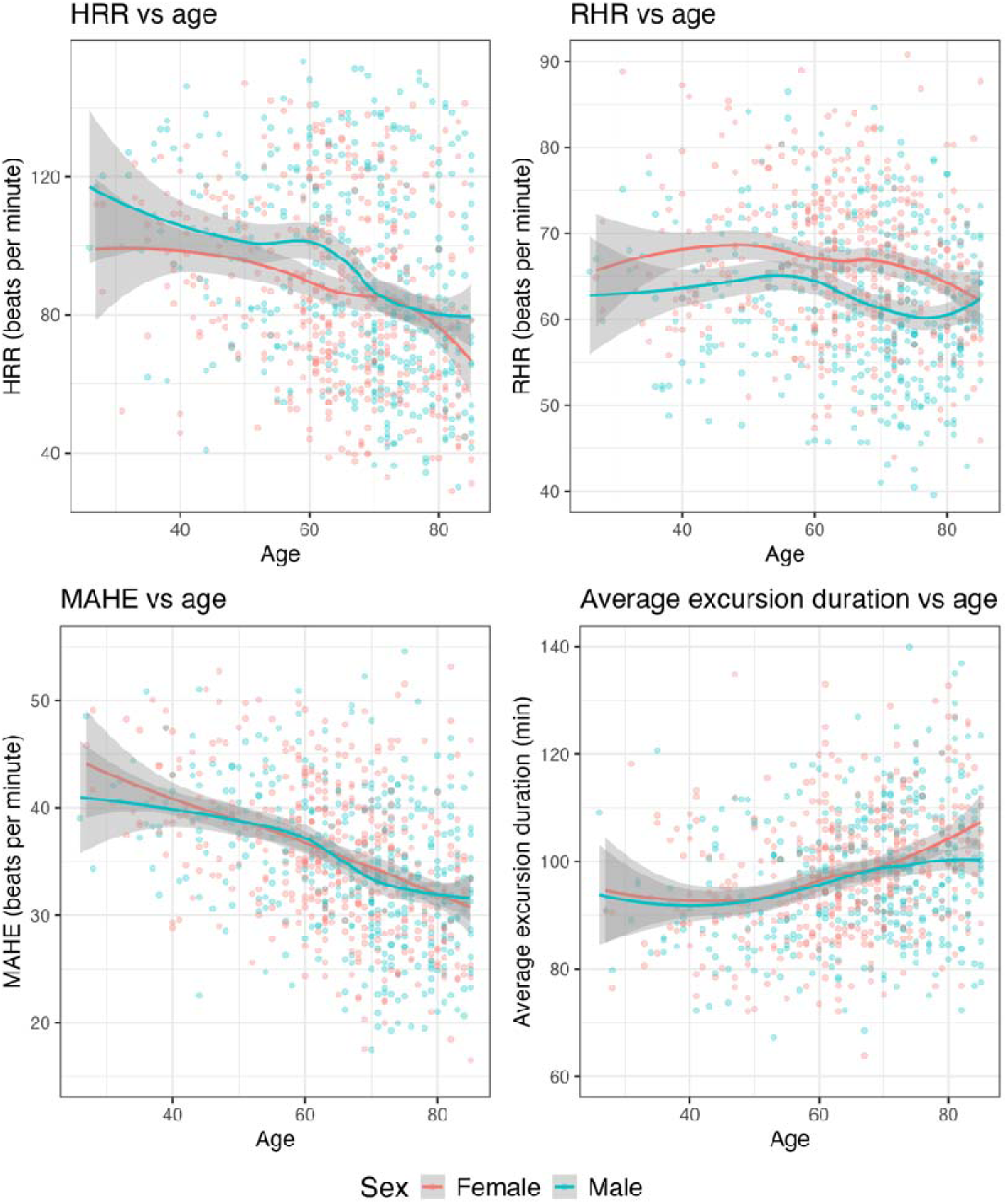
Age trends in HR summary metrics. Scatterplots with fitted local polynomial regression curves for HRR, RHR, MAHE, and average HRE duration as functions of age. Red represents females and blue represents males. Axis limits were selected to emphasize the central age trends; observations outside the displayed range were retained in model fitting.

Conventional HRR was lower with increasing age in both sexes. At age 30, fitted HRR was approximately 120 bpm in males and 100 bpm in females, whereas by age 85 it was approximately 80 bpm in both groups. In contrast, RHR remained comparatively stable across age. Females generally had higher RHR than males, while males showed only a modest late-life reduction.

The HRE summaries provided additional information not contained in conventional HRR. MAHE decreased from approximately 40–45 bpm at age 30 to around 30 bpm by age 85 in both sexes, indicating smaller amplitudes among the qualifying free-living HR events detected by the algorithm. Average excursion duration was approximately 90 minutes through midlife and increased after age 55. The increase was somewhat more pronounced among older females.

Thus, excursion amplitude and excursion duration changed in different directions with age. For the representative ages 40 and 80 used in Figure 5, fitted mean excursion amplitude decreased from approximately 40 to 32 bpm, while fitted mean excursion duration increased from approximately 90 to 104 minutes. Older adults tended to have smaller but more prolonged detected HR excursions.

### Analysis of Diurnal HR Curves

We used functional data analysis (FDA), which extends conventional regression models to outcomes represented as continuous functions.^17^ Function-on-scalar regression (FoSR) allows subject-specific curves such as dHR and dHRR to be modeled as outcomes while scalar characteristics such as age, sex, and BMI enter as predictors.

For participant *i*, the dHR model can be written as

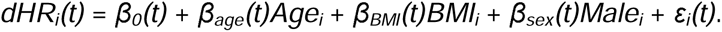

Similarly, the dHRR model is

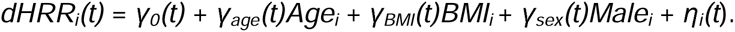

Age and BMI were centered by their mean, and females were used as the reference group. Because the regression coefficients are functions of time, associations are allowed to vary over the 24-hour cycle. For example, the association between age and dHR can be substantially stronger in the afternoon than overnight rather than being constrained to one average age effect. Figure 4 shows fitted dHR and dHRR curves from the two FoSR models.

**Figure 4.**
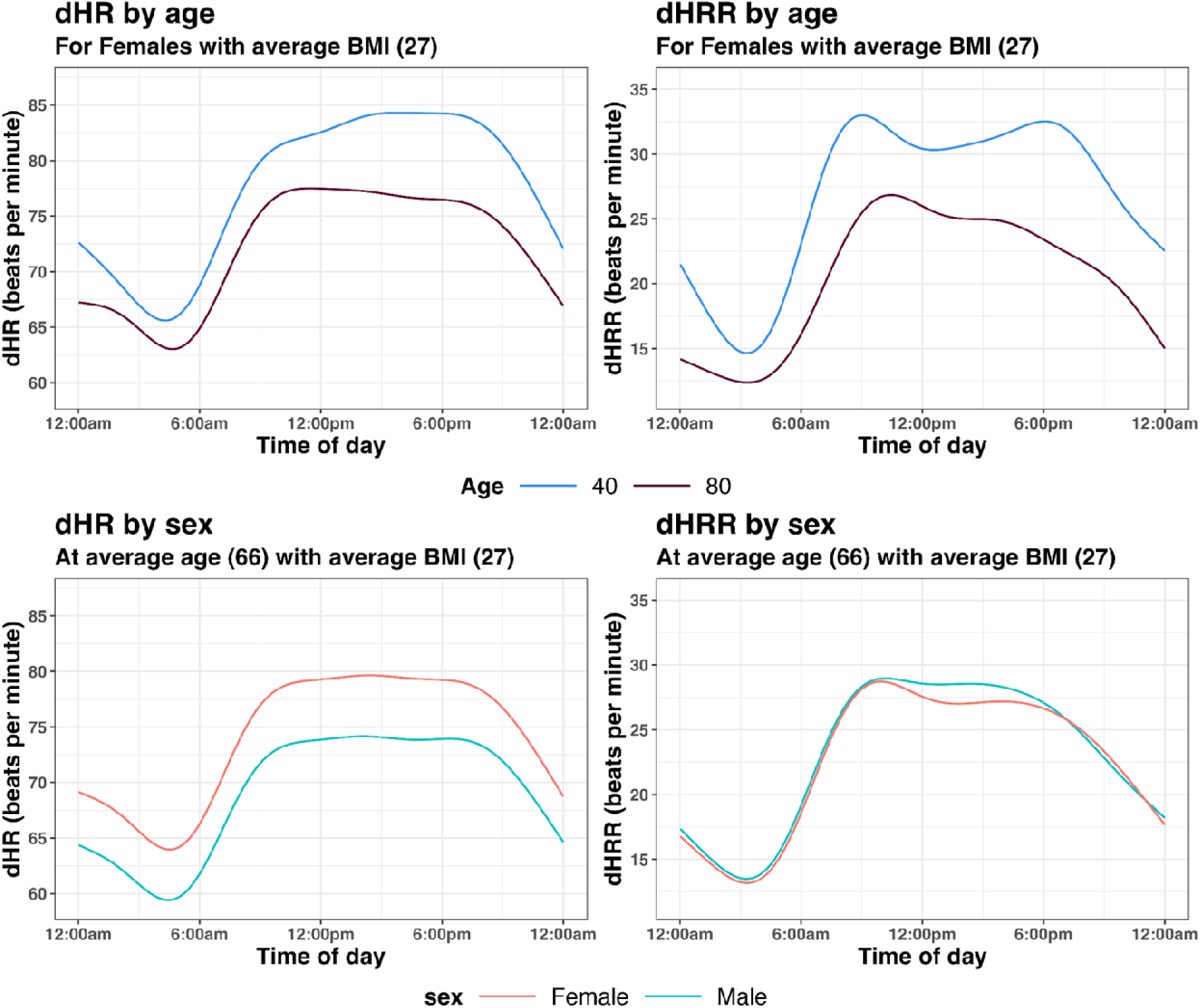
Fitted dHR and dHRR curves from function-on-scalar regression models. Left: fitted median dHR. Right: fitted dHRR. Top panels show age-specific fitted curves for females with mean BMI; bottom panels show sex-specific curves at mean age and BMI.

#### Diurnal Heart Rate

dHR decreased with increasing age across the entire 24-hour period, but the magnitude of this association was strongly time dependent. For all age groups, dHR was lowest in the early morning, increased sharply during the morning hours, remained relatively high during the daytime, and declined during the evening and night. The age-related difference was strongly time dependent; between ages 40 and 80, fitted dHR was approximately 2 bpm lower at 3:00 AM but 8 bpm lower at 6:00 PM (Supplementary Figure 2). Among participants at the mean age and BMI, females had consistently higher dHR than males across the whole 24-hour period.

This result illustrates one advantage of treating wearable HR as a function: a scalar average could show lower HR with age but would not indicate when during the day the age difference is greatest.

#### Diurnal Heart Rate Range

The dHRR analysis revealed a second layer of age-related information. At younger ages, dHRR increased sharply during the morning, with a peak around 9:00 AM, followed by a modest decline and a second later-day peak around 6:00 PM. With increasing age, the dHRR envelope became narrower across much of the day. In addition, its temporal organization changed. The later-day peak decreased progressively and was largely absent by approximately age 70. At the same time, the morning peak shifted later, approaching noon at the oldest ages.

Thus, conventional HRR and dHRR provided related but non-equivalent information. Conventional HRR showed that the overall observed HR span becomes smaller with age. dHRR additionally showed where this contraction occurs and how the timing and shape of the available daily range are reorganized.

Sex differences in dHRR were comparatively small, with only modest time-localized differences. Supplementary Figures 4 and 5 show the estimated coefficient functions and pointwise 95% confidence bands for the dHR and dHRR models. Supplementary Figure 3 displays fitted curves according to BMI.

**Figure 5.**
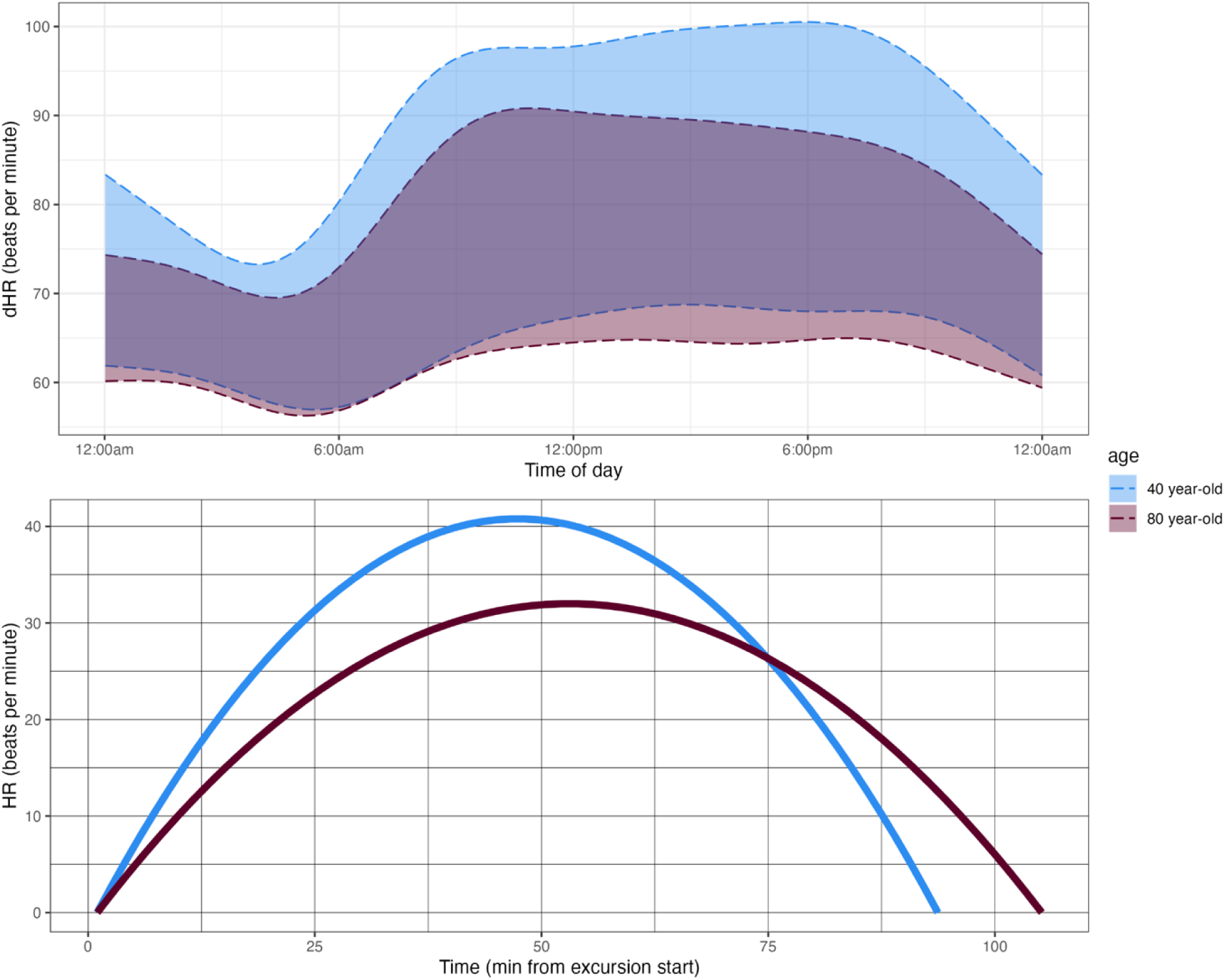
Age differences in dHRR and heart rate excursions. Top: fitted diurnal HR envelopes for representative ages 40 and 80 years derived from the dHR and dHRR models. Bottom: illustrative excursion reconstruction using age-specific fitted MAHE and average HRE duration. The lower panel is a visualization of fitted feature values and is not an observed exercise-recovery trajectory.

### Integrating Diurnal Range and Excursion Dynamics

Figure 5 illustrates age-related differences in HR dynamics at complementary scales. The upper panel compares fitted dHR envelopes for representative 40- and 80-year-old females with BMI 27 kg/m². Envelope width represents fitted dHRR and illustrates how HR range changes across the day: at age 80, the range is narrower, particularly during waking hours, and its diurnal pattern is reorganized relative to age 40.

To illustrate these fitted age trends using a common contrast, the lower panel compares HRE characteristics at ages 40 and 80. Mean excursion amplitude decreases from approximately 40 to 32 bpm, while mean excursion duration increases from approximately 90 to 104 minutes. For visualization, HREs are reconstructed from the fitted mean amplitude and duration from the local polynomial regressions in Figure 3, with a parabola interpolated between these values. These reconstructed curves combine two fitted HRE features and should not be interpreted as average physiological trajectories or estimates of post-exercise HR recovery.

Together, these representations show distinct and complementary features of age-related HR dynamics: a smaller global HR range, compression and reorganization of HR range across the day, and smaller, longer-lasting individual HR excursions.

## 5. Discussion

This study demonstrates that continuous wearable HR data reveal complementary features of HR variability that are obscured by conventional scalar summaries. In BLSA, aging was associated with a smaller global HR range, compression and reorganization of the diurnal HR range across the day, and smaller, longer-lasting individual HR excursions. These findings illustrate the complementary information provided by conventional HRR, dHRR, and HREs at global, diurnal, and event levels.

dHRR extends conventional HRR by showing how the observed HR range is organized across the 24-hour day. In BLSA, the age-related contraction in global HRR was not uniform across clock time: the later-day peak was attenuated, and the morning expansion occurred later at older ages. Importantly, dHRR represents the range expressed during free-living monitoring rather than maximal physiological capacity. It therefore reflects both cardiovascular responsiveness and the activities and exposures encountered during habitual life. This distinction separates dHRR from standardized exercise testing, which measures response to a controlled challenge. Prior wearable studies have shown that resting HR has a strong 24-hour rhythm and that variability in repeated daily RHR contains information beyond mean RHR.^16,24^ However, these studies characterize HR level or variability as separate scalar summaries. dHRR extends these concepts by describing how the repeatedly observed HR range itself varies across clock time. The age-related decline in dHR closely paralleled the established diurnal pattern of autonomic modulation. In more than 8 million Fitbit users, HRV peaked in the early morning (∼3am) and reached its nadir in the evening (∼6pm),^25^ suggesting that the age-related attenuation of diurnal HR may reflect diminished diurnal autonomic modulation across the daily cycle.

HREs revealed a complementary age-related pattern that was not apparent from global or diurnal range alone. Between ages 40 and 80, mean excursion amplitude decreased while mean excursion duration increased, indicating two parallel changes in the characteristics of individual HR events. The distinction between amplitude and duration is particularly important because large, brief excursions and smaller, prolonged excursions may produce similar aggregate summaries while representing different temporal patterns. One interpretation is that aging leads to smaller HR responses that take longer to return toward baseline, consistent with known age-related changes in cardiovascular and autonomic function.^26^ Consistent with this, a global wearable-HR analysis in over five million adults found within-person cardiac responses to environmental stressors strongly blunted at older ages, indicating reduced autonomic reserve.^27^ The decline in HRE amplitude observed in our study may represent a related event-level, free-living pattern. Note that our specific implementation of HREs captures HR dynamics at an intermediate temporal scale, coarser than beat to beat fluctuations summarized by HRV but finer than the 24-hour diurnal organization captured by dHRR. HREs therefore complement, rather than substitute for, conventional autonomic HRV indices.

The proposed measures also fit within the broader effort to derive digital biomarkers of biological aging from wearables,^28^ where circadian and HRV features have been tied to aging and longevity.^29^ Excursions that are simultaneously smaller and longer-lasting describe responses that are both blunted and slower to resolve, and a slowing of recovery dynamics with age has been proposed as a feature of declining physiological resilience.^30^ Whether HRE duration tracks this resilience loss longitudinally is a natural question for future work.

Standardized and open-source implementation may facilitate reproducibility and comparison across studies.^31^ The *ihr* package makes the proposed measures, algorithmic choices, and visualizations directly available for evaluation and extension, while the AHP provides a common participant-level representation linking the raw HR signal with diurnal and conventional summaries. The primary contribution of this study is the proposed analytical framework; the BLSA analysis demonstrates the additional structure dHR and HRE reveal rather than establishing these characteristics as biomarkers of cardiovascular aging. If validated, measures that localize HR-range contraction to particular times of day, or that separate the amplitude and persistence of free-living HR responses, could provide complementary digital markers of autonomic and chronotropic function than global summaries, with potential relevance to conditions such as heart failure and chronotropic incompetence.^20^

## 6. Limitations and Future Directions

Free-living dHRR and HREs reflect both cardiovascular response characteristics and the exposures encountered during monitoring. A participant who performs little vigorous activity may have a narrow observed range despite preserved maximal exercise capacity. This is a particular concern for the age associations reported here, because older adults tend to be less physically active, and part of the observed narrowing of dHRR and dampening of HRE amplitude may therefore reflect lower habitual activity rather than reduced cardiovascular responsiveness.^32^ A parallel concern applies to the timing results. Because dHR and dHRR are pooled by clock time, the later morning peak observed at older ages may in part reflect differences in the timing of activity and sleep rather than temporal reorganization of the daily HR envelope. The Actiheart records accelerometry concurrently with HR, so activity intensity can be incorporated directly into the function-on-scalar regression models. Future analyses should incorporate concurrent activity and environmental conditions when evaluating these age associations. Free-living HR and its variability are also systematically modulated by ambient temperature, season, and latitude.^27^ Because BLSA monitoring is distributed across participants and times of year, some age differences in dHRR and HRE features could reflect environmental conditions of monitoring, which future work should adjust for alongside activity.

Both proposed measures also depend on analytic choices that have not yet been evaluated for HR. The HRE detection procedure is adapted from MAGE methods developed for CGM.^18,19^ The moving-average approach provides a transparent way to identify peaks and nadirs of HREs while suppressing high-frequency noise, but optimal window lengths and amplitude thresholds for HR remain to be established. Varying the short and long smoothing windows and the standard-deviation threshold, and comparing a fixed absolute amplitude threshold, would establish whether the observed decrease in excursion amplitude and increase in excursion duration with age are robust. dHRR raises a related estimation concern: it is computed from repeated measurements at each clock time, and with relatively few valid days, extreme quantiles may be sensitive to individual observations and to the smoothing procedure. Future work should examine the stability of dHRR as a function of monitoring duration and its sensitivity to alternative quantile ranges such as 10th-to-90th percentile bands. The temporal scale of HREs is determined by the detector’s smoothing windows and amplitude threshold, an adjustable analytic choice rather than an intrinsic property of the signal. Different settings may identify excursions at finer or coarser scales, while the retained minute level information allows additional features, including frequency, area, rise and decline slopes, event spacing, and time of occurrence, to be examined across these alternative resolutions. Characterizing HR dynamics across different temporal scales and assessing their sensitivity to detector settings are important directions for future work.

We also note that the age trends in landmark and excursion measures (Figure 3) are summarized with fitted local-polynomial curves without accompanying confidence bands or formal tests, so the reported amplitude and duration differences with age should be regarded as descriptive; the function-on-scalar models, by contrast, report pointwise 95% confidence bands (Supplementary Figures 4–5). Two definitional choices also warrant caution: RHR is computed over a fixed 3:00–7:00 AM window that presumes sleep or rest during that interval, which may not hold for individuals with atypical or shifted schedules and could interact with the diurnal timing results; and Actiheart HR is itself derived through device-specific processing of the ECG signal, which may influence excursion detection independently of the detection algorithm studied here.

This BLSA analysis is cross-sectional and based on a relatively healthy, well-characterized aging cohort in which the oldest participants are health-selected survivors. Beyond limiting generalizability, this selection can distort the apparent shape of the age curves themselves, because the individuals contributing at the oldest ages are not representative of their birth cohorts. The observed age associations should therefore be interpreted as an illustration of the framework rather than as longitudinal estimates of within-person aging. Adding the repeated visits from BLSA in future work would allow assessment of whether the cross-sectional patterns are also observed within individuals as they age.

The motivating large-scale cohorts, including All of Us and ABCD, predominantly use wrist-worn photoplethysmography (PPG), whereas the present demonstration uses a research-grade chest-worn electrocardiogram monitor. PPG-derived HR is subject to different noise characteristics, motion artifact, and missingness,^33^ which could affect both dHRR estimation and HRE detection. The measures introduced here will therefore require separate validation on consumer PPG data, and consistent computation across studies, cohorts, and devices, before results can be compared across settings.

Finally, the clinical and prognostic value of these measures remains to be established. The present study demonstrates that they retain information not present in conventional summaries, but it does not show that dHRR or HRE features improve prediction of cardiovascular events or other clinical outcomes. Linking HREs to concurrent activity and to standardized exercise testing would clarify how free-living response magnitude and persistence relate to chronotropic competence, fitness, autonomic regulation, and formal HR recovery, which is an important step for interpreting these features as measures of cardiac reserve.^34^

## Data and Code Availability

The *ihr* R package implementing the measures described here is openly available at https://github.com/IrinaStatsLab/ihr. Analysis code to reproduce the reported results is available from the corresponding author on reasonable request. BLSA data are available through the National Institute on Aging Intramural Research Program subject to approval.

