## Supplemental Materials for "Diurnal Heart Rate Range and Heart Rate Excursions: Novel Characterizations of Wearable Heart Rate Variability With an Application to Aging"

**Supplementary Materials**

**Open-Source *ihr* Package and Its Functionality**

This section provides implementation detail for the open-source Interpreting Heart Rate (*ihr*) R package introduced in the main text, focusing on excursion detection, HR episodes, and rate-of-change measures. Conventional summaries, diurnal quantile curves, dHRR, and the Ambulatory Heart Rate Profile are described in the main text.

**Heart Rate Excursions and MAHE**

The implementation of HRE detection in *ihr* is adapted from the open-source moving-average algorithm developed by Fernandes et al. for calculating Mean Amplitude of Glycemic Excursions in CGM data.^19^ Short and long moving averages represent two levels of temporal smoothing. Crossings of the moving-average curves identify intervals likely to contain local maxima or minima. Peaks and nadirs are subsequently identified within these intervals. Candidate excursion amplitudes are filtered so that only rises and declines meeting the specified standard-deviation threshold are retained as qualifying events. In the current implementation, MAHE is computed as the mean amplitude of qualifying HREs. Importantly, the software retains the event segmentation itself. Consequently, the framework is not limited to MAHE. HRE duration, event timing, rise and decline time, frequency, and distributions of these properties can also be calculated. The smoothing parameters and excursion thresholds were adapted from CGM methodology and should not be interpreted as established HR-specific physiological cutoffs. Future validation should determine how parameter choices affect HRE reliability and their relationships with concurrent activity and standardized physiological measures.

**Episode Calculation and Rate of Change**

Episodes provide an additional representation of localized HR behavior. An episode is defined as a contiguous interval during which HR reaches a specified participant-specific activity threshold and remains above that level for a prespecified duration.

For example, if a participant's threshold for light physical activity is 80 bpm and HR exceeds 80 bpm for an extended period, the corresponding interval can be classified as a light-activity HR episode. Episodes preserve information on duration, timing, intensity, and frequency that is lost when HR is summarized solely as total time in an activity category. Episodes and HREs are related but answer different questions. An episode is defined relative to an external or participant-specific HR threshold, whereas an HRE is defined by the local structure of rises and declines in the HR time series. The package also calculates minute-to-minute rate of change (RoC),

*RoC(t)* = [*HR(t)* − *HR(t − Δt)*] / *Δt*.

For minute-level observations, *Δt* is one minute. The resulting sequence of RoC values characterizes short-term changes in HR. Larger absolute values indicate more rapid local changes. *ihr* supports visualization of the RoC distribution and calculation of its standard deviation as an overall scalar measure of short-term HR change. These additional metrics are included as extensions of the broader package. The primary methodological focus of the present paper remains dHRR and HREs.

**Supplementary Figures**

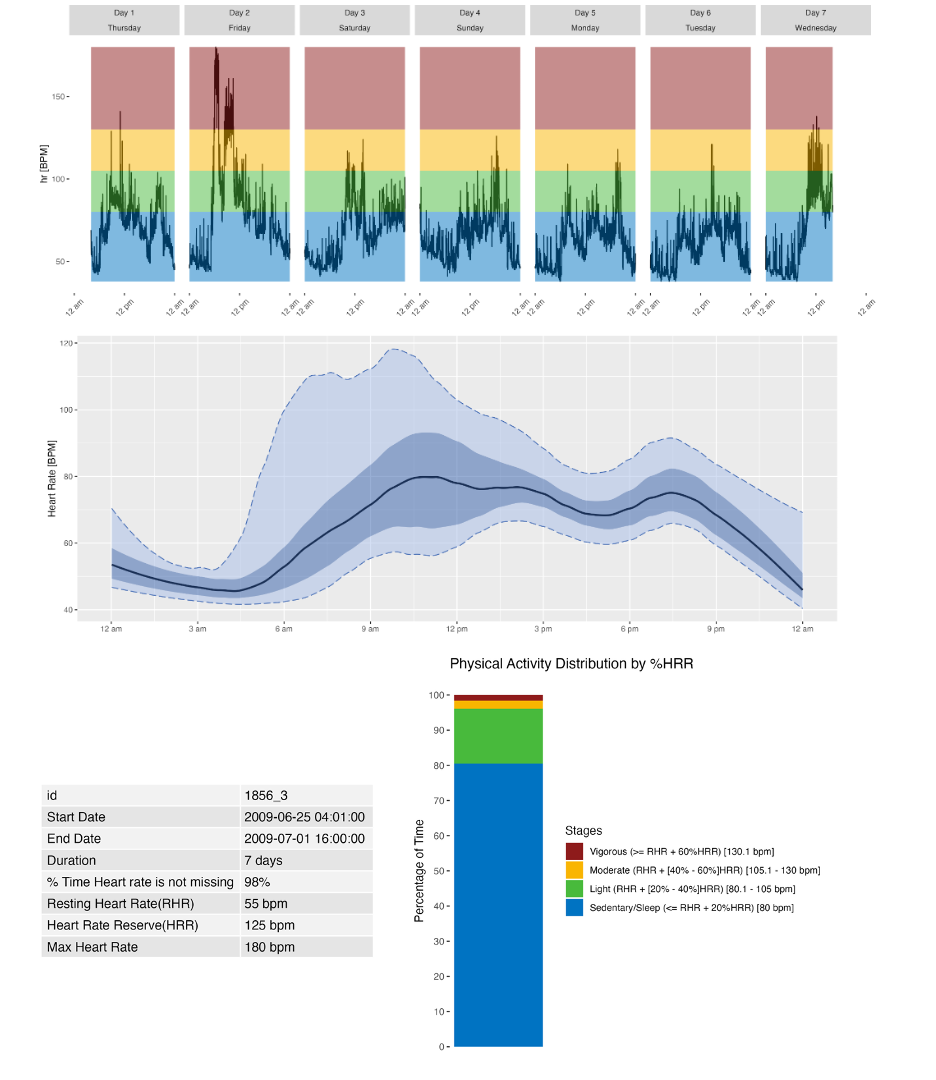

**Supplementary Figure 1.** Ambulatory Heart Rate Profile (AHP) for one subject in BLSA. Top panel: Seven days of minute-level HR data. X-axis shows time of day, y-axis shows HR in beats per minute, and each window represents a day of wear. Colors show physical activity intensity threshold derived from the subject’s HRR.

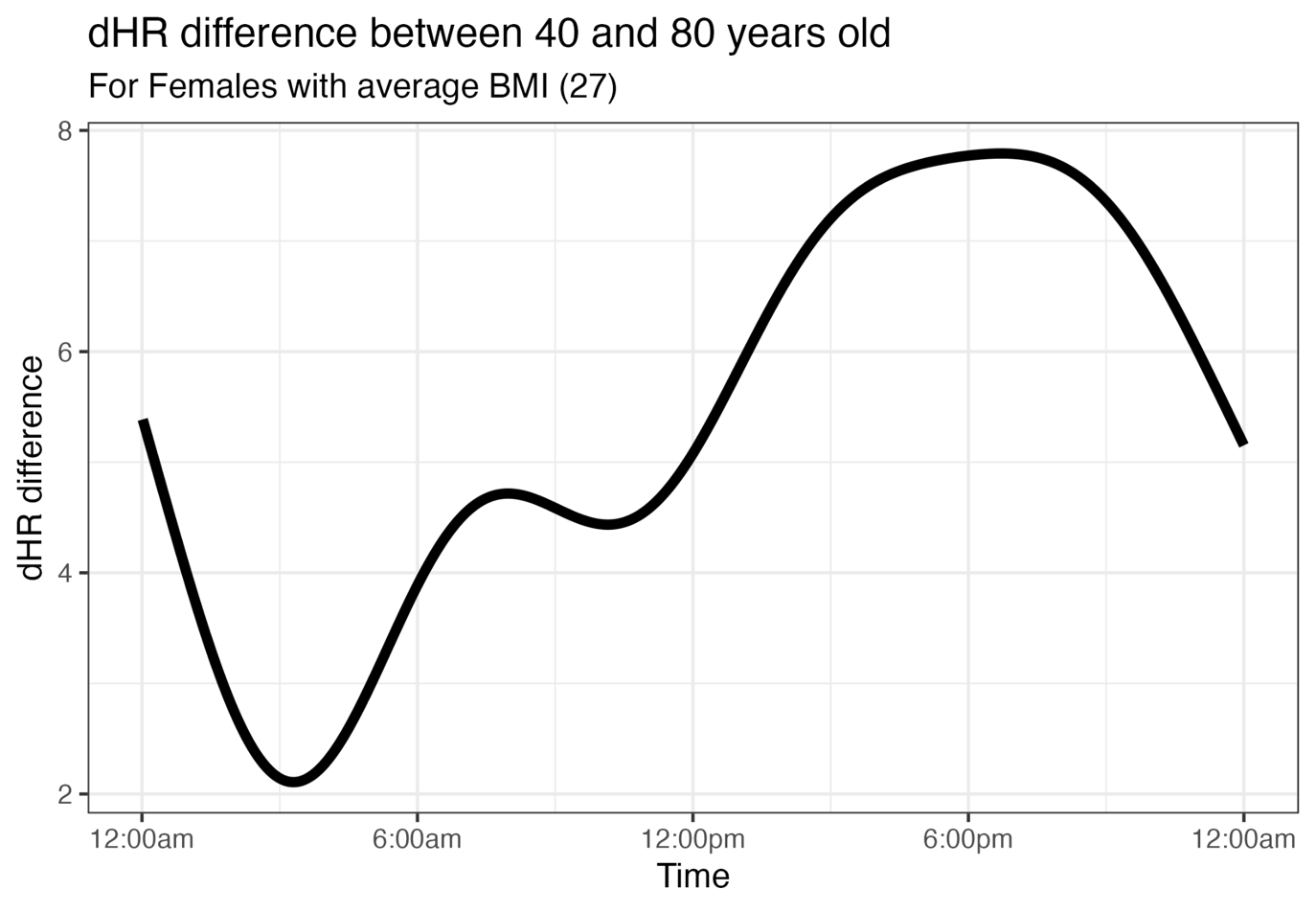

**Supplementary Figure 2.** Age-related differences in diurnal heart rate at ages 40 and 80 years. The contrast between fitted dHR curves at ages 40 and 80 shows that the age-related difference varies substantially over the 24-hour cycle, with the largest reduction occurring in the late afternoon and early evening and the smallest difference during the overnight period.

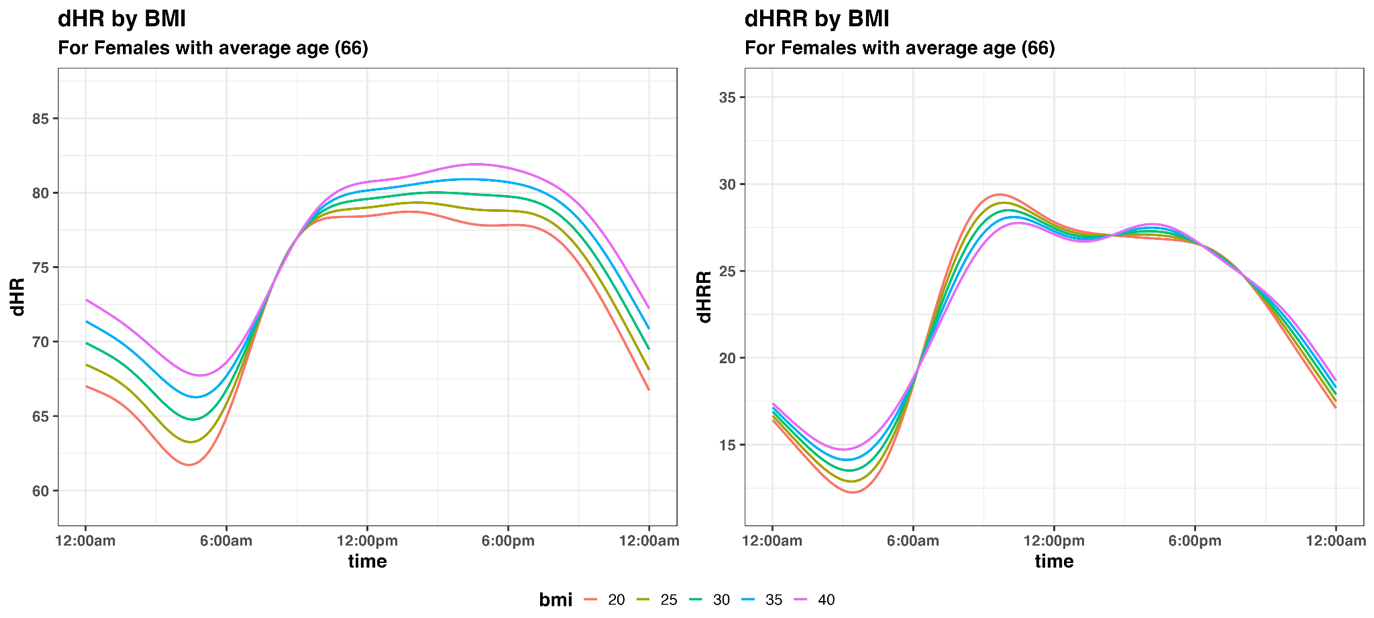

**Supplementary Figure 3.** Fitted diurnal heart rate and diurnal heart rate range curves by BMI. Left: fitted dHR according to BMI. Right: fitted dHRR according to BMI. Differences in dHR are more apparent than differences in dHRR, which remains relatively similar across BMI levels for much of the 24-hour period.

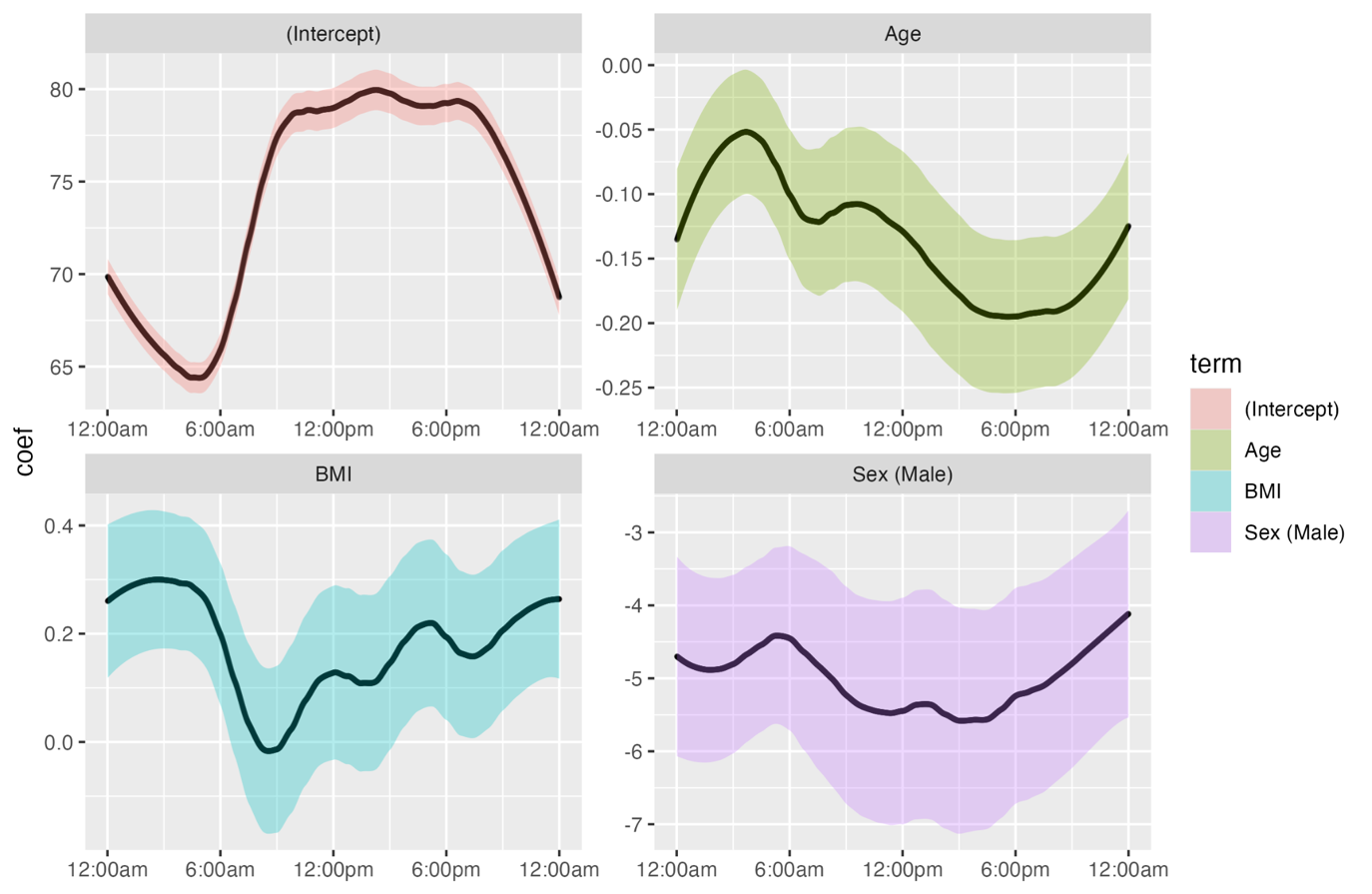

**Supplementary Figure 4.** Estimated coefficient functions from the function-on-scalar regression model for diurnal heart rate. Estimated functions are shown for the intercept, age, BMI, and sex, with pointwise 95% confidence bands.

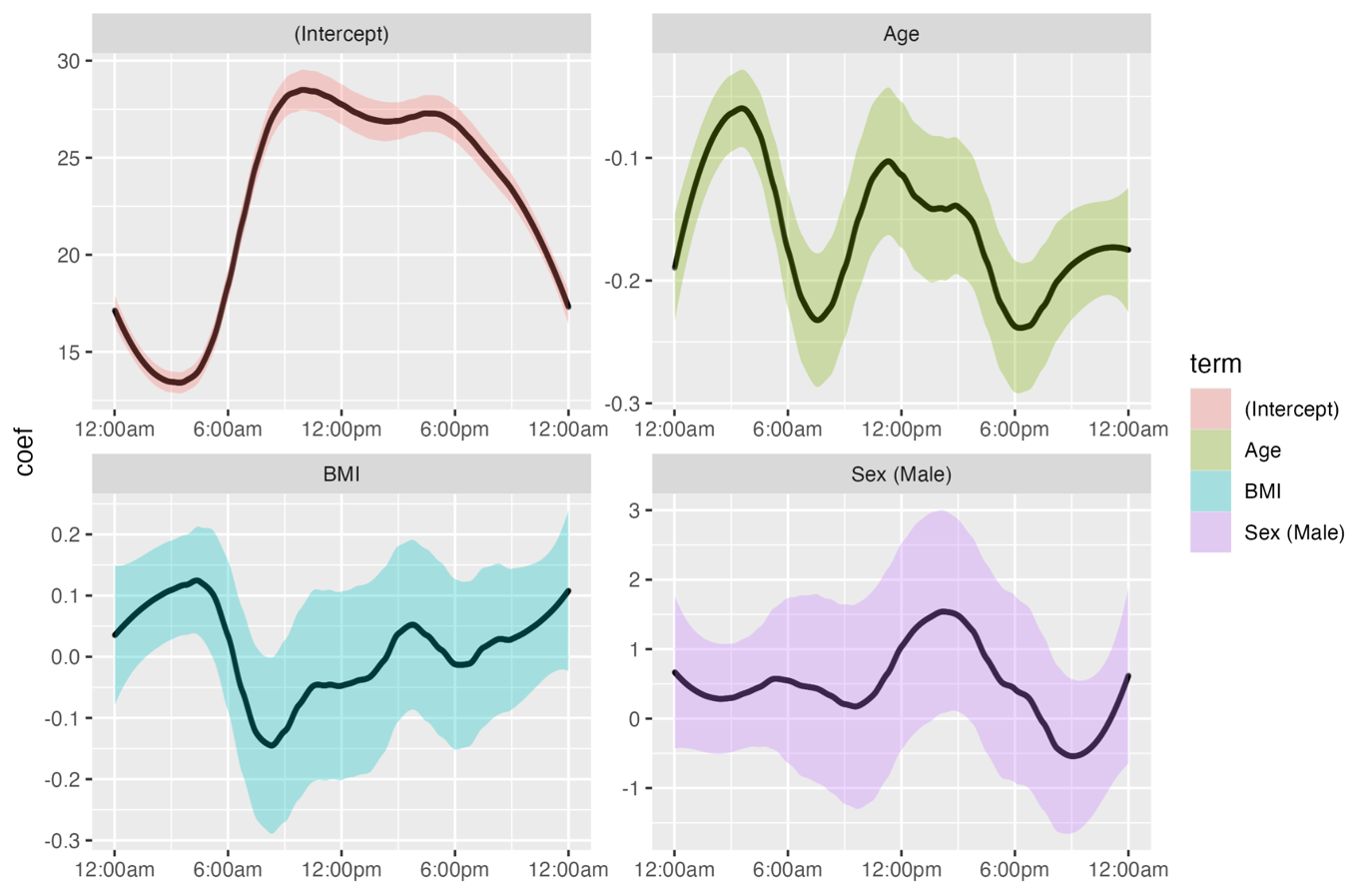

**Supplementary Figure 5.** Estimated coefficient functions from the function-on-scalar regression model for diurnal heart rate range. Estimated functions are shown for the intercept, age, BMI, and sex, with pointwise 95% confidence bands.
